# Comparison of excess deaths and laboratory-confirmed COVID-19 deaths during a large Omicron epidemic in 2022 in Hong Kong

**DOI:** 10.1101/2024.07.04.24309940

**Authors:** Hualei Xin, Alexandra H. T. Law, Justin K. Cheung, Yun Lin, Peng Wu, Zhongjie Li, Benjamin J. Cowling, Weizhong Yang, Jessica Y. Wong

## Abstract

**Background:** Using a local elimination strategy, Hong Kong was able to minimize COVID-19 mortality in 2020 and 2021, but a large epidemic caused by the Omicron variant occurred in 2022. We aimed to estimate the overall, age-, sex-, epidemic period- and cause-specific excess mortality in 2022 in Hong Kong and compared excess mortality to laboratory-confirmed COVID-19 mortality.

**Methods:** Negative binomial regression analysis was used to model time series of weekly all-cause and cause-specific deaths from 2010 to 2021 to predict the weekly number of deaths in 2022 against counterfactual baselines projected from the trends in the absence of a pandemic. The estimated excess deaths were compared with laboratory-confirmed COVID-19 deaths overall and by age and epidemic period.

**Results:** We estimated that there were 13,500 (95% CI: 13,400-13,600) excess deaths in 2022, which was slightly higher than the 12,228 deaths recorded with laboratory-confirmed COVID-19, with the majority of the excess deaths and laboratory-confirmed deaths occurring among older adults. The increased number of excess deaths over laboratory-confirmed COVID-19 deaths was most substantial from February to April 2022 (a difference of 847 deaths), when the largest Omicron wave peaked. Most of the excess deaths (78%) were from respiratory causes, while 10% were from cardiovascular causes. A slight reduction in malignant neoplasm mortality was identified among older adults in 2022.

**Conclusions:** A substantial increase in population mortality was identified in 2022 in Hong Kong, slightly larger than the laboratory-confirmed COVID-19 deaths. Apart from the possibility of underdiagnosis, excess deaths might also be attributed to the overload of healthcare resources during the pandemic. Deaths from COVID-19 may have displaced some deaths that would otherwise have occurred due to other causes although we did not find evidence of substantial mortality displacement.

## INTRODUCTION

Since the emergence of SARS-CoV-2 at the end of 2019, millions of COVID-19 deaths with infection of the virus have been recorded globally (1). However, the reported number may not represent the full mortality impact associated with the COVID-19 pandemic for several reasons. First, some COVID-19 deaths may be assigned to other causes due to misclassification or misdiagnosis, particularly during the early phase of the pandemic when the diagnostic capacity was more limited (2, 3). Second, there may have been indirect health impact due to the disruption of essential medical services due to pressure on the health-system (3), changes in medical behavior resulting from difficulty in accessing healthcare services (4, 5), or fear of getting infected in medical institutions or delays in seeking healthcare under stay-at-home policies or other social distancing measures (5, 6). Excess mortality is the difference between the observed and expected mortality obtained under some counterfactual scenario (7, 8). This indicator has long been used to estimate the disease burden of past health crises or pandemics based on mortality time-series (8–10). For the COVID-19 pandemic, the World Health Organization (WHO) estimated 14.83 million excess deaths in 2020 and 2021, which is 2.74 times higher than the reported deaths (5.42 million) from laboratory-confirmed SARS-CoV-2 infections (3).

Hong Kong adopted an elimination strategy during the COVID-19 pandemic in 2020 and 2021 (11). Individual and population interventions were introduced, such as universal isolation of confirmed cases, strict quarantine of close contacts, school closures and other compulsory social distancing measures (12). These measures effectively controlled pre-Omicron transmission resulting in 181 (2 per 100,000) local laboratory-confirmed COVID-19 deaths in 2020 and 30 (0.4 per 100,000) in 2021 (12). However, our previous study estimated an estimated 1,800 excess deaths from diseases other than COVID-19 in 2020, potentially related to delays or reductions in healthcare seeking as a consequence of strict public health and social measures (5). In early 2022 Hong Kong experienced a very large Omicron epidemic in the early period of the year, with over 2 million confirmed COVID-19 cases (by RT-PCR or rapid antigen tests) and 10,000 deaths reported (136 per 100,000) (12).

The considerable excess mortality identified during the elimination phase with minimal transmission of COVID-19 (5) prompted us to investigate mortality impact of COVID-19 over a much larger epidemic caused by Omicron subvariants in Hong Kong by estimating the excess mortality in 2022 by age, sex, epidemic period, and cause of death, and comparing the excess mortality to the reported fatal cases with confirmed COVID-19.

## METHODS

### Sources of data

Individual death data from 2010 to 2022 were obtained from the Census and Statistics Department of Hong Kong. Weekly mortality data were aggregated into groups of age (0-4, 5-14, 15-44, 45-64, 65-79 and ≥80), sex, and cause of death. Cause of deaths were coded by the International Classification of Diseases Tenth Revision (ICD-10) and grouped into: all causes (A00-Z99), and the following major causes: respiratory diseases (J00-J99), malignant neoplasms (C00-C97), cardiovascular diseases (I00-I99), kidney diseases (N00-N07, N17-N19, N25-N27), external causes (S00-T88), diabetes mellitus (E10-E14), and other causes (including diseases of the digestive system, mental and behavioral disorders, certain infectious and parasitic diseases, diseases of the nervous system, et al.). Data on individual patients with laboratory-confirmation of SARS-CoV-2 in 2022 were obtained from the Hong Kong Hospital Authority (HA), Centre for Health Protection and Deaths Registry (13). Epidemiological information of individual patients was provided in the database, including age, sex, date of laboratory confirmation, occurrence of death in-hospital or out-of-hospital. In this analysis, a laboratory-confirmed COVID-19 death was defined as a person who had a positive RT-qPCR result testing for SARS-CoV-2 with a respiratory specimen collected within 28 days before or seven days after the date of death. The annual mid-year population data by age and sex between 2010 and 2022 were collected from the Census and Statistics Department (14).

### Statistical analysis

Weekly mortality risk was calculated by dividing the number of weekly deaths by mid-year populations in Hong Kong, by age, sex, and cause of death. We applied negative binomial regression models to the time series of weekly number of all-cause and disease-specific deaths from 2010 to 2021, assuming the temporal mortality in 2010-21 extending to 2022. In each regression model, we included the population size as an offset and accounted for long-term trends using the calendar year. Fourier terms with the variable “week” were included in the regression models to allow for annual seasonality. Another covariate was included in the model to account for changes in the mortality trend in Hong Kong since the emergence of SARS-CoV-2 in January 2020. The COVID-19-associated excess mortality risk was calculated by subtracting the predicted weekly mortality risk from the fitted regression model from the observed weekly mortality risk. Performance of the model was examined using the Akaike Information Criterion (AIC) during the model proceeding.

In 2022, Hong Kong experienced two major epidemics driven by Omicron subvariants, including the largest wave during February - April and a smaller wave from November onwards. We defined weeks 1-26 as period 1 (wave 5), weeks 27-45 as period 2 (wave 6a), and weeks 46-52 as period 3 (wave 6b) (12). The all-cause excess mortality was estimated for these three periods using the same method described above and compared with the reported COVID-19 mortality for each period. Further technical details of the statistical methods are provided in the Appendix. All statistical analyses were conducted in R, version 4.1.0 (R Foundation for Statistical Computing, Vienna, Austria).

Our study received ethical approval from the Institutional Review Board of the University of Hong Kong/Hospital Authority Hong Kong West Cluster.

## RESULTS

An overall increasing trend of annual mortality in Hong Kong was observed from 2010 through to 2022, with a slight increase from 596 per 100,000 population in 2010 to 645 in 2019, and a sharp rise to 864 in 2022 (Figure 1A). Prior to 2022, the weekly mortality peaked in late December to January every year while a prominent peak was noted in February - March in 2022, in line with the trend in confirmed COVID-19 case numbers (Figure 1A, Figure 1B). In 2022, people aged 80 years or above had the highest mortality, followed by individuals aged 65-79 years, and the mortality risk was generally higher in males than females. The top three causes of death in 2022 were respiratory diseases (302 per 100,000 population), malignant neoplasms (202), and cardiovascular diseases (152), which differed from 2020 and 2021 when malignant neoplasms (197 and 203 in 2020 and 2021, respectively) were the leading cause, followed by respiratory (151 and 157) and cardiovascular (137 and 140) diseases (Table S1).

**Figure 1.**
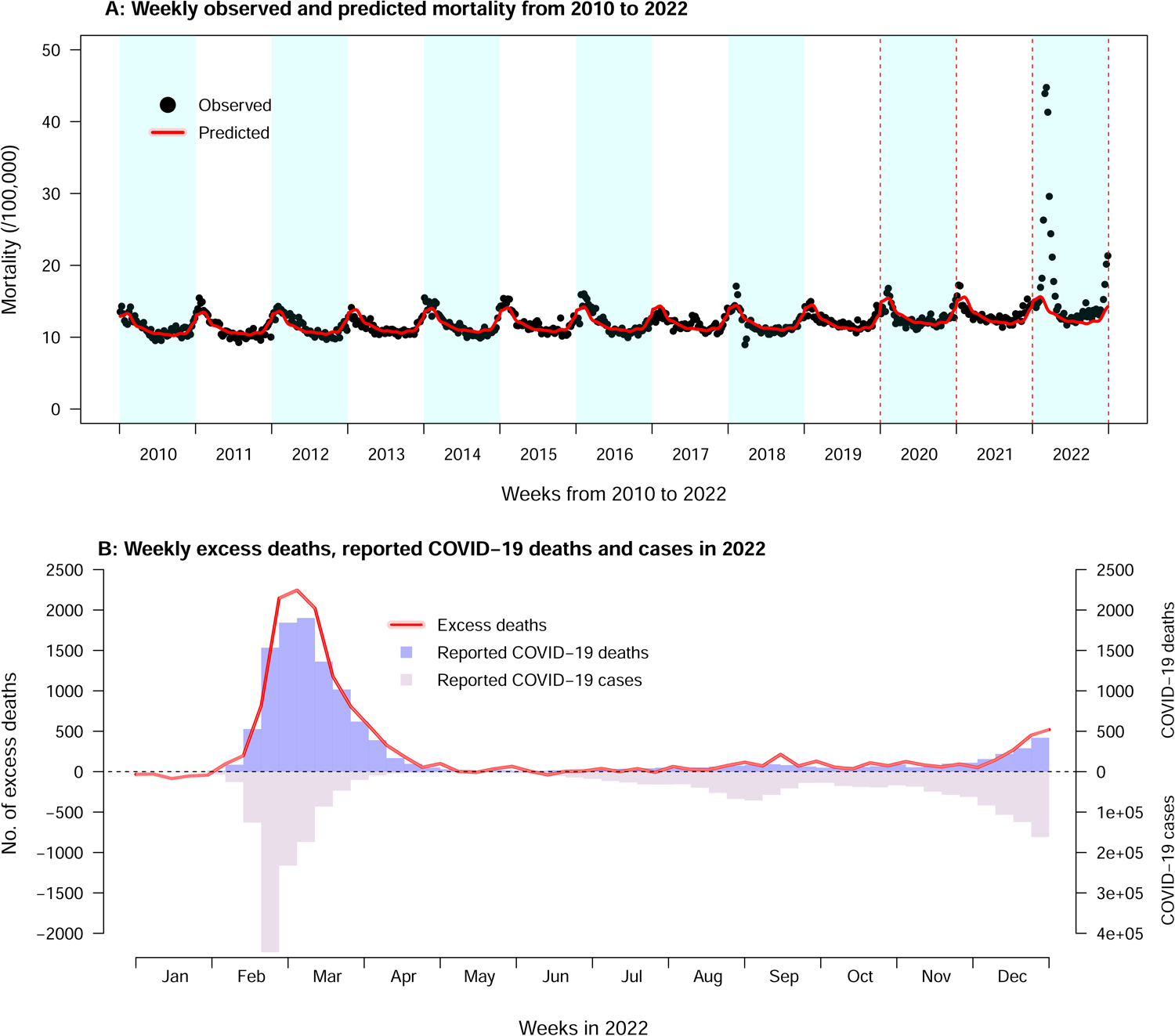
The overall trend of mortality and excess deaths in 2022 in Hong Kong. **A**) Weekly observed (black dots) and predicted (red solid line and the pink shaded area show the mean predicted weekly mortality and 95% confidence intervals, respectively) mortality from 2010 to 2022 in Hong Kong. **B**) Predicted weekly numbers of excess deaths (red solid line and the pink shaded area show the mean weekly numbers of excess deaths and 95% CI, respectively), weekly numbers of reported COVID-19 deaths (purple bars) and weekly numbers of locally infected COVID-19 cases in Hong Kong in 2022.

In total, there were 63,484 deaths (864 per 100,000 population) recorded by the death registry in 2022. With the predicted counterfactual mortality baseline of 49,966 deaths (680, 95% CI: 678, 682, per 100,000 population), the excess mortality in 2022 was estimated to be 184 per 100,000 (95% CI: 182, 186), equating to an estimated 13,518 (95% CI: 13,403-13,633) additional deaths and a 27% increase in all-cause mortality (Figures 1A and 2A, Table S2). People aged 80 years or above showed the highest excess mortality risk in 2022 (2,354 per 100,000 population, 95% CI: 2,330-2,378) throughout the three defined epidemic periods, followed by 65-74 years (260, 95% CI: 255-265), accounting for 68% and 22% of all excess deaths in this year, respectively (Figure 2, Table S2). Overall males (232, 95% CI: 230-235 per 100,000) showed a higher excess mortality than females (143, 142–145) and sex differences were indicated in most age groups, particularly people over 80 years (2,897 excess deaths, 95% CI: 2,858-2,935 for males vs 1,960, 1,930-1,991 for females). Less than 2% of the estimated excess deaths were identified in individuals younger than 45 years (Figure S1A, Table S2).

**Figure 2.**
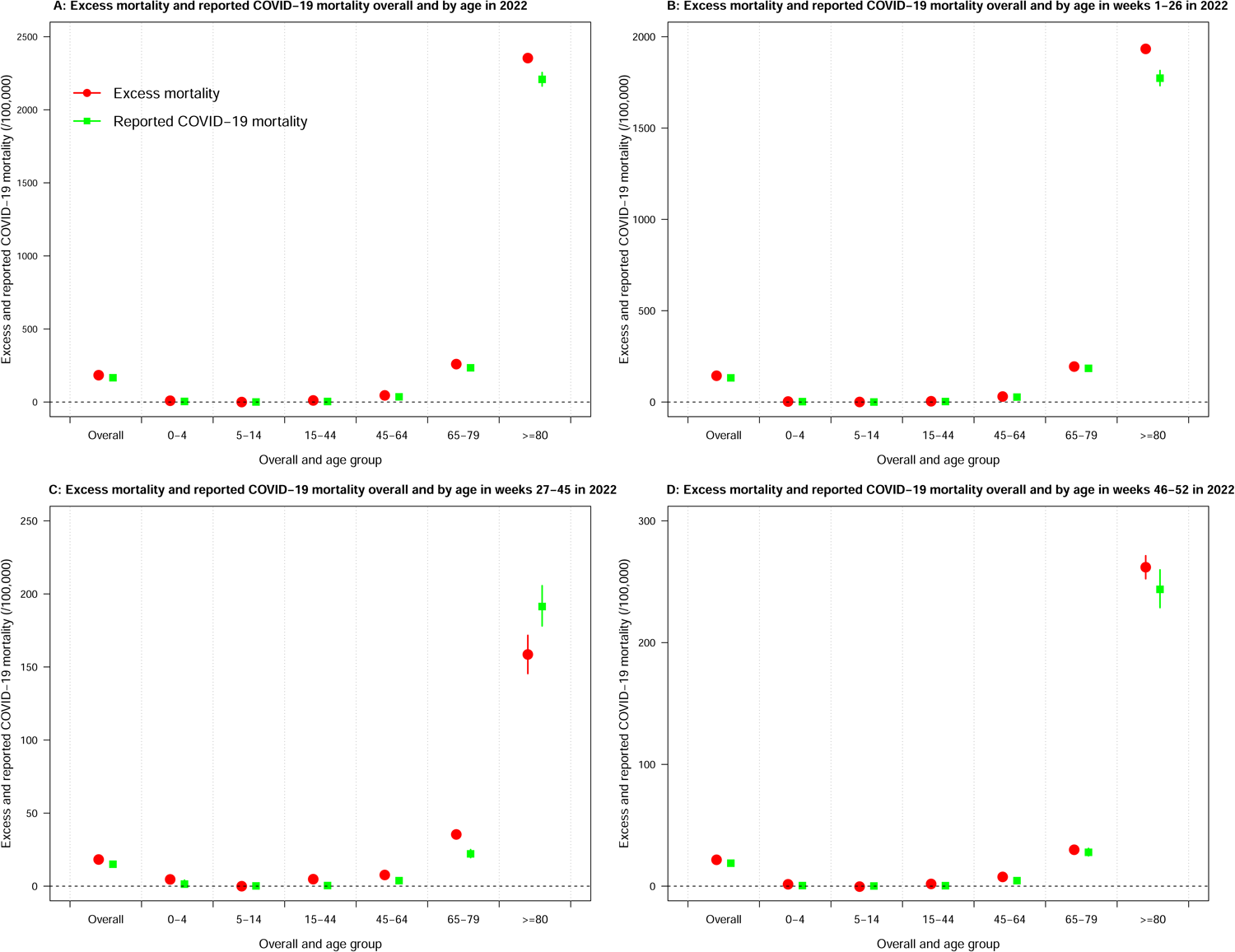
Overall and age-specific excess mortality and reported COVID-19 mortality in 2022. **A)** Comparison of excess mortality and reported COVID-19 mortality overall and by age in 2022. **B)** Comparison of excess mortality and reported COVID-19 mortality overall and by age in weeks 1-26 in 2022. **C)** Comparison of excess mortality and reported COVID-19 mortality overall and by age in weeks 27-45 in 2022. **D)** Comparison of excess mortality and reported COVID-19 mortality overall and by age in weeks 46-52 in 2022.

Compared with 12,228 individuals died with confirmed COVID-19 recorded in 2022 (166 deaths per 100,000 population, 95% CI: 164-169), the slightly higher estimate of the excess mortality (184 per 100,000 population, 95% CI: 182-186) for the same year shared similar age-, sex-, and period-specific patterns (Figure 2A). Older adults over 80 years of age contributed to most of the differences between excess deaths and the reported COVID-19 deaths (44%, 567/1290 deaths increased), followed by those aged 65-79 years (287, 22%) and 45-64 years (236, 18%) (Figure 2A, Table S3). The excess mortality was also higher than the reported COVID-19 mortality for both males (232 vs 212 per 100,000 population) and females (143 vs 128). The sex- and age-specific analysis indicated that 63% of the overall difference between the excess and reported deaths in females was attributed to patients over 80 years of age. However, the death difference in males was contributed by the four adult groups, 15-44 (20%), 45-64 (30%), 65-79 (23%), and those over 80 years (27%) similarly (Figure S2, Figure S3).

Among the 7 causes of death investigated in this study, excess mortality was identified in 2022 for these major causes except for malignant neoplasms (Figure 3, Figure S4). The highest excess mortality risk was estimated for respiratory diseases (145 per 100,000 population, 95% CI: 144-146) and cardiovascular diseases (17, 95% CI: 16-18), representing 92% and 13% increase from the estimated baseline, respectively. The additional deaths from the two causes accounted for 88% of all excess deaths estimated for 2022 (Figure 3, Table S2). The reported mortality from malignant neoplasms (202 per 100,000 population) was slightly lower than the predicted (205, 95% CI: 204-207), with the largest reduction in people over 80 years (reduced by 25 per 100,000 population, 95% CI: 18-32), followed by those 65-79 years (6, 95% CI: 3-9) (Figure 3B, Table S2).

**Figure 3.**
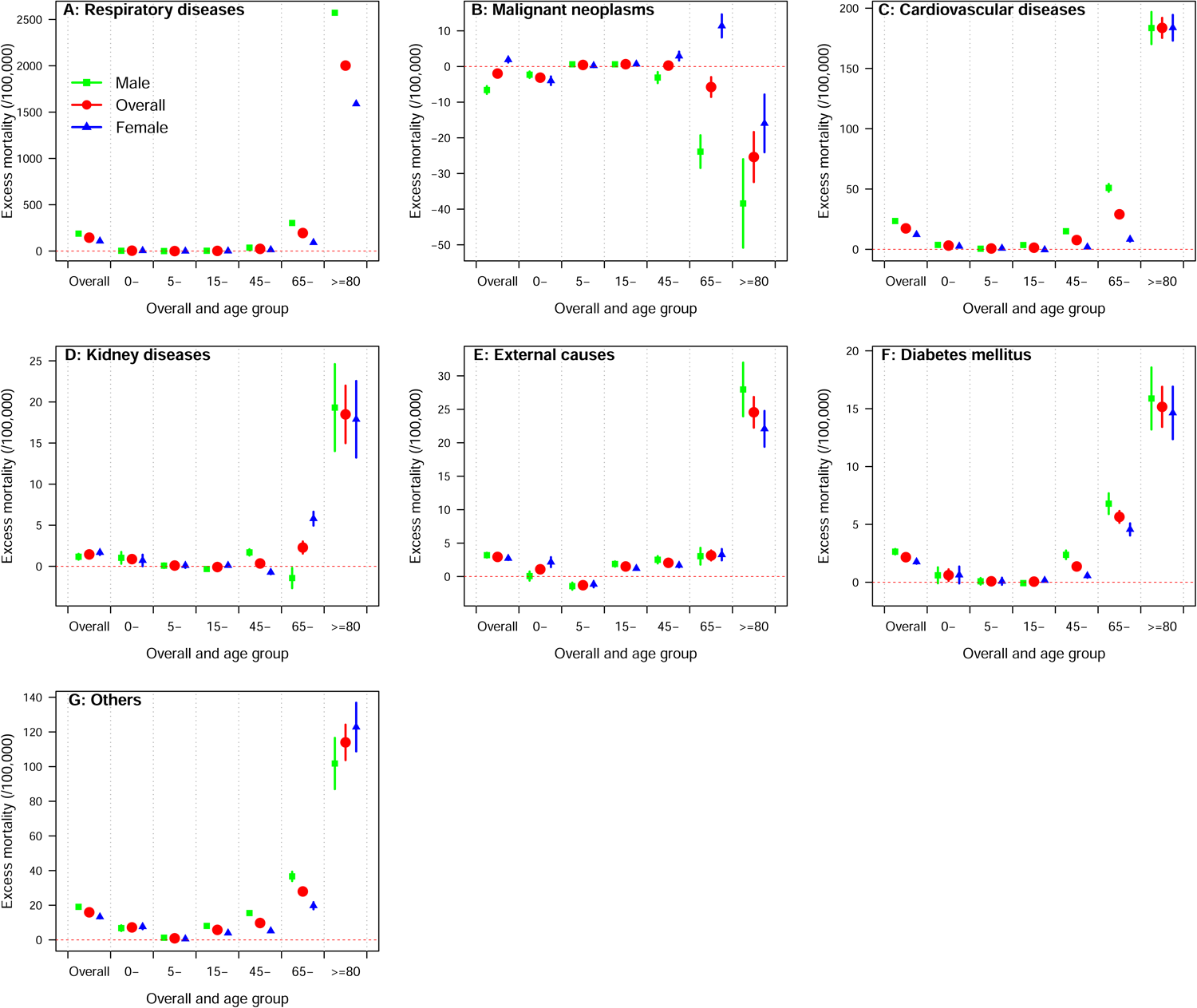
Cause-specific excess mortality by age and sex in Hong Kong in 2022. A) Excess mortality by age and sex for respiratory diseases. B) Malignant neoplasms. C) Cardiovascular diseases. D) Kidney diseases. E) External causes. F) Diabetes mellitus. G) Others (deaths from all other causes except those causes shown in panels A-F).

The higher number of excess deaths over reported COVID-19 deaths in 2022 was most prominent in period 1 (weeks 1-26, 847 more deaths, 95% CI: 762-932), accounting for about 66% (847/1290) of the overall difference (Figure 2B-D, Table S3). Among these, the majority occurred in older adults over 80 years of age (624/847,74%) (Figure 2B). During period 2, however, the estimated excess deaths in people over 80 years were lower than the reported COVID-19 deaths (−128, 95% CI: −179, −77) although the age group 65-79 years showed the highest increase in the number of excess deaths over the reported COVID-19 deaths (151, 120-183) among all age groups, followed by 15-44 years (116, 109-123) (Figure 2C, Table S3). In period 3, people aged over 80 (over 71, 34-107) and 45-64 (over 73, 60-86) showed a similar number of increases (Figure 2D, Table S3). The increased number of excess deaths over confirmed COVID-19 deaths was identified in all epidemic periods for both males and females (Figure S2, Figure S3).

## DISCUSSION

We estimated that the excess mortality associated with the COVID-19 pandemic in 2022 was 184 (95% CI: 182, 186) per 100,000 population, 27% higher than the baseline mortality predicted for the year based on the historical trend. The estimated excess mortality was not only identified in causes of respiratory diseases, but also other causes, such as cardiovascular diseases, accounting for 10% of the overall excess deaths in 2022, particularly among older adults at age of 80 years or above. A slightly higher number of excess deaths than the reported COVID-19 deaths was observed mainly in the older adults during the peak of the Omicron BA.2 wave in early 2022.

A close concordance was indicated in our study between the estimated number of excess deaths and the officially recorded deaths with a confirmation of COVID-19 in 2022. Globally, a report from the World Health Organization found that the excess mortality in 2020 and 2021 was 2.74 times higher than the reported COVID-19 mortality (3). A series of excess mortality studies conducted in the US found that among the overall excess deaths estimated during the pandemic, only 65%-72% were attributed to COVID-19 (15–18). Hong Kong had very substantial laboratory testing capacity, testing approximately 3% of the population each day during March 2022 which included routine on-admission testing in hospitals (12). Nevertheless in our previous analysis of population mortality in Hong Kong in 2020 we found an increase in cardiovascular mortality with very few COVID-19 deaths, potentially attributable to changes in healthcare seeking behaviours or disruption in care for older individuals with cardiovascular diseases (5), and this has also been reported in other locations (15–19). In the present analysis we did not observe a substantial indirect impact of the pandemic on mortality from other causes, although some indirect impact likely occurred and is included as part of the overall excess mortality.

One notable difference between the substantial mortality impact of COVID-19 in Hong Kong compared with other locations is that the impact in Hong Kong occurred in 2022, whereas highest excess mortality rates in the pandemic in other parts of the world generally occurred in 2020 and early 2021 prior to the availability of COVID-19 vaccines (3, 21, 22). We have reported separately that vaccination uptake in older adults in Hong Kong was low for various reasons (12, 23, 24), leading to a substantial mortality impact in the Omicron BA.2 wave in March-April 2022 (25). We also reported that case fatality rates increased by 3-fold during the peak weeks of the Omicron wave compared to earlier and later in that epidemic (26), likely due to the extreme pressure on healthcare resources at that time.

The higher number of excess deaths than the deaths with confirmed COVID-19 in Hong Kong in 2022 identified mainly in older adults during the peak of the Omicron wave in this study might also be the consequence of the large number of infections and the relatively lower vaccination coverage in the elderly (12). In contrast, countries such as Canada and Norway estimated a lower excess all-cause mortality during the Omicron epidemic than the reported COVID-19 mortality (18). It could be explained that many laboratory-confirmed COVID-19 deaths particularly in very frail older adults were actually deaths “with COVID-19” rather than “due to COVID-19”, which were counted as COVID-19 deaths but not changing the overall mortality rate (2, 20). Consequently, the death rate from causes other than COVID-19 may be reduced due to mortality displacement, and this might explain the reduction we observed in deaths from malignant neoplasms in the Hong Kong population throughout 2022 as well as the relatively lower excess deaths in older adults over months after the largest omicron wave in comparison with the confirmed COVID-19 deaths.

A number of limitations exist in this study. First, we were not able to separate the direct impact from indirect impact of the COVID-19 pandemic in the estimated excess deaths. Previously Lee et al. intended to achieve the separation by including covariates representing the two types of impact in the regression model (19). Future studies are needed to clearly define the different impact and develop valid methods to make a causal inference on the health impact caused by COVID-19. Second, the accuracy of the estimated excess mortality for some defined groups in the analysis, such as children aged 5-14 years, might be limited by the small numbers of weekly deaths. Third, weekly death data from 2010 to 2021 was used to obtain the predicted weekly deaths in 2022. Different from previous studies which often used the data before 2020 as the mortality baseline, our baseline was determined based on the historical mortality trend as well as allowing for possible changes during the COVID-19 pandemic. Therefore, a direct comparison of our estimates with others may not be entirely appropriate.

In conclusion, a substantial number of excess deaths were identified in Hong Kong during a large Omicron predominant COVID-19 epidemic, and the slightly higher number of excess deaths over reported laboratory-confirmed COVID-19 deaths demonstrated the importance of pandemic responses with effective pharmaceuticals in particular vaccines in reducing the overall health burden especially in older adults. We did not identify substantial mortality displacement from other causes of death in 2022.

## Data Availability

Restrictions apply to the availability of these data. The mortality data are available for access with the permission from the Hospital Authority and the Census and Statistics Department of Hong Kong.

## ACKNOWLEDGMENTS

The authors thank Julie Au for technical support.

## AUTHOR CONTRIBUTIONS

All authors meet the ICMJE criteria for authorship. The study was conceived by BJC, PW and WY. HX, JYW, AHTL and JKC analyzed the data. HX wrote the first draft of the manuscript. All authors provided critical review and revision of the text and approved the final version.

