## Appendix for "Comparison of excess deaths and laboratory-confirmed COVID-19 deaths during a large Omicron epidemic in 2022 in Hong Kong"

*Additional information on statistical analysis*

The estimated excess mortality in 2022 in this study was the difference between the observed mortality in the study period and the mortality that would have been expected in that period, based on historical data from 2010 to 2021. The interrupted time-series regression models were used throughout the study to obtain the overall and subgroup excess mortality. We then created a dataset comprising weekly aggregated deaths from 2010 to 2022 for each unique combination of age group and sex (referred to as age-sex stratum). In each age-sex stratum, we fitted a negative binomial regression model using the weekly number of deaths from 2010 to 2021, accounting for long-term trends, seasonality, and the impact of the COVID-19 pandemic by including a dummy variable with the value of 1 if the calendar weeks were during 2020 and 2022 (COVID-19 period) and 0 for weeks outside of the period to accound for possible changes in mortality in Hong Kong since the emergency of SARS-CoV-2 at the start of 2020, and using the population in Hong Kong as an offset. Then, the obtained model was used to predict the weekly number of deaths with standard error (SE) in 2022 in that stratum. Models were screened according to the Akaike Information Criterion (AIC) obtained during the model proceeding. In the final selected model, we used a Fourier term with eight harmonics within one year and the variable "week" to indicate the seasonality, the variable "year" as an indicator of long-term trend, and a dummy variable with the value 1 if the weeks were during 2020 and 2022 (COVID-19 period) and 0 for other weeks to indicate the possible changing trend of mortality after the occurrence of the COVID-19 pandemic. Finally, we summed the predicted number of deaths across the age and sex strata to obtain the predicted counterfactual baseline deaths in 2022, and the overall excess deaths were estimated as the difference between the observed and predicted deaths in all age groups. The age-, sex-, or epidemic period-specific excess deaths were calculated by summing the predicted number of deaths in the corresponding stratum. The estimated excess mortality was compared with the reported laboratory-confirmed COVID-19 mortality overall and by age, sex, and epidemic period.

The overall and subgroup excess deaths/mortality uncertainties were estimated based on the standard error of the predicted number of deaths, i.e., the square root of the sum of the variances (squared standard errors) of the predicted weekly number of deaths from the regression models in 2022. For mortality of the reported COVID-19 deaths, a binomial test was used to obtain their 95% confidence intervals (95% CIs).

### Table S1. Characteristics of the number of deaths and mortality risk recorded by the death resgistry between 2010 and 2022 in Hong Kong.

| Characteristics | 2010 | 2011 | 2012 | 2013 | 2014 | 2015 | 2016 | 2017 | 2018 | 2019 | 2020 | 2021 | 2022 |
|---|---|---|---|---|---|---|---|---|---|---|---|---|---|
| **Overall** | 41392 (589) | 41395 (585) | 42864 (599) | 42573 (592) | 44338 (612) | 45417 (623) | 45875 (625) | 45826 (620) | 45768 (614) | 47977 (639) | 49479 (661) | 50875 (686) | 63484 (864) |
| **Period** | | | | | | | | | | | | | |
| Weeks 1-26 | 21590 (307) | 22333 (316) | 23329 (326) | 21939 (305) | 23999 (331) | 24376 (334) | 24815 (338) | 23380 (316) | 23900 (321) | 24716 (329) | 25193 (337) | 25980 (350) | 36848 (502) |
| Weeks 27-45 | 14235 (203) | 13628 (193) | 13941 (195) | 14654 (204) | 14455 (200) | 15112 (207) | 15025 (205) | 16192 (219) | 15476 (208) | 16430 (219) | 17315 (231) | 17517 (236) | 18223 (248) |
| Weeks 46-52 | 5567 (79) | 5434 (77) | 5594 (78) | 5980 (83) | 5884 (81) | 5929 (81) | 6035 (82) | 6254 (85) | 6392 (86) | 6831 (91) | 6971 (93) | 7378 (100) | 8413 (115) |
| **Sex** | | | | | | | | | | | | | |
| Male | 22928 (696) | 23105 (700) | 23805 (715) | 23607 (709) | 24452 (731) | 24918 (740) | 25484 (755) | 25313 (746) | 25385 (744) | 26635 (778) | 27209 (796) | 28006 (828) | 35436 (1056) |
| Female | 18464 (495) | 18290 (485) | 19059 (498) | 18966 (492) | 19886 (510) | 20499 (522) | 20391 (515) | 20513 (513) | 20383 (504) | 21342 (523) | 22270 (548) | 22869 (567) | 28048 (703) |
| **Age** | | | | | | | | | | | | | |
| 0- | 171 (72) | 152 (61) | 152 (58) | 130 (50) | 132 (50) | 77 (27) | 124 (44) | 111 (40) | 97 (35) | 62 (22) | 98 (40) | 76 (33) | 86 (40) |
| 5- | 49 (8) | 45 (8) | 50 (9) | 50 (9) | 44 (8) | 55 (10) | 34 (6) | 45 (8) | 40 (7) | 54 (9) | 43 (7) | 55 (10) | 47 (8) |
| 15- | 1449 (47) | 1391 (45) | 1383 (44) | 1373 (44) | 1257 (41) | 1347 (44) | 1264 (42) | 1221 (41) | 1209 (41) | 1201 (41) | 1078 (38) | 1129 (41) | 1328 (50) |
| 45- | 6946 (322) | 7159 (324) | 7108 (316) | 7224 (318) | 7356 (320) | 7464 (323) | 7545 (324) | 7449 (318) | 7258 (307) | 7652 (321) | 7547 (312) | 7618 (317) | 8537 (359) |
| 65- | 12627 (1902) | 12078 (1803) | 12284 (1769) | 11741 (1634) | 11840 (1585) | 12061 (1534) | 12215 (1484) | 12136 (1411) | 11954 (1331) | 12704 (1348) | 13335 (1343) | 13839 (1305) | 17134 (1504) |
| 80- | 20150 (7911) | 20570 (7576) | 21887 (7666) | 22055 (7315) | 23709 (7486) | 24413 (7436) | 24693 (7256) | 24864 (7014) | 25210 (6852) | 26304 (6928) | 27378 (7053) | 28158 (7194) | 36352 (9338) |
| **Causes** | | | | | | | | | | | | | |
| Respiratory diseases | 8430 (120) | 8696 (123) | 9529 (133) | 9216 (128) | 9759 (135) | 10440 (143) | 10622 (145) | 10354 (140) | 10439 (140) | 11456 (153) | 11269 (151) | 11656 (157) | 22155 (302) |
| Malignant neoplasms | 12934 (184) | 13131 (186) | 13288 (186) | 13486 (188) | 13648 (188) | 14173 (194) | 14133 (193) | 14359 (194) | 14395 (193) | 14767 (197) | 14705 (197) | 15018 (203) | 14848 (202) |
| Cardiovascular diseases | 10462 (149) | 10204 (144) | 10129 (142) | 9768 (136) | 10157 (140) | 9859 (135) | 9935 (135) | 9957 (135) | 9291 (125) | 9651 (129) | 10268 (137) | 10402 (140) | 11176 (152) |
| Kidney diseases | 1462 (21) | 1525 (22) | 1626 (23) | 1567 (22) | 1684 (23) | 1644 (23) | 1679 (23) | 1678 (23) | 1581 (21) | 1655 (22) | 1714 (23) | 1771 (24) | 1821 (25) |
| External causes | 1532 (22) | 1417 (20) | 1403 (20) | 1514 (21) | 1386 (19) | 1534 (21) | 1451 (20) | 1492 (20) | 1428 (19) | 1562 (21) | 1513 (20) | 1535 (21) | 1723 (23) |
| Diabetes mellitus | 523 (7) | 448 (6) | 384 (5) | 353 (5) | 386 (5) | 482 (7) | 494 (7) | 407 (6) | 464 (6) | 493 (7) | 579 (8) | 546 (7) | 712 (10) |
| Others | 6049 (86) | 5974 (84) | 6505 (91) | 6669 (93) | 7318 (101) | 7285 (100) | 7561 (103) | 7579 (103) | 8170 (110) | 8393 (112) | 9431 (126) | 9947 (134) | 11049 (150) |

**Table S2. Characteristics of excess mortality estimated in 2022 in Hong Kong.**

| Characteristics | Model-obtained mortality baseline (/100,000) | Excess mortality (/100,000) | Proportion of excess mortality to baseline mortality (%) | No. of excess deaths (95% CI) | Proportion (% of overall excess deaths) |
|---|---|---|---|---|---|
| **Overall** | 680 | 184 (182, 186) | 27 | 13518 (13402, 13633) | 100 |
| **Period** | | | | | |
| Weeks 1-26 | 358 | 144 (143, 145) | 40 | 10585 (10500, 10670) | 78 |
| Weeks 27-45 | 230 | 18 (17, 19) | 8 | 1341 (1278, 1404) | 10 |
| Weeks 46-52 | 93 | 22 (21, 22) | 24 | 1592 (1547, 1637) | 12 |
| **Sex** | | | | | |
| Male | 824 | 232 (230, 235) | 28 | 7791 (7707, 7874) | 58 |
| Female | 559 | 143 (142, 145) | 26 | 5727 (5648, 5806) | 42 |
| **Age** | | | | | |
| 0- | 31 | 9 (8, 10) | 29 | 19 17, 22 | 0 |
| 5- | 8 | 0 (0, 1) | 0 | 0 (-2, 3) | 0 |
| 15- | 39 | 11 (11, 11) | 28 | 293 (282, 305) | 2 |
| 45- | 314 | 45 (44, 47) | 14 | 1082 (1048, 1115) | 8 |
| 65- | 1244 | 260 (255, 265) | 21 | 2959 (2903, 3015) | 22 |
| 80- | 6984 | 2354 (2330, 2378) | 34 | 9164 (9070, 9258) | 68 |
| **Causes** | | | | | |
| Respiratory diseases | 158 | 145 (144, 145) | 92 | 10634 (10581, 10687) | 79 |
| Malignant neoplasms | 205 | -2 (-3, -1) | - | -145 (-193, -97) | -1 |
| Cardiovascular diseases | 136 | 17 (17, 18) | 13 | 1275 (1232, 1317) | 9 |
| Kidney diseases | 24 | 1 (1, 2) | 4 | 106 (89, 123) | 1 |
| External causes | 21 | 3 (3, 3) | 14 | 216 (200, 231) | 2 |
| Diabetes mellitus | 8 | 2 (2, 2) | 25 | 159 (149, 169) | 1 |
| Others | 135 | 16 (15, 17) | 12 | 1169 (1122, 1215) | 9 |

**Table S3. Excess deaths and reported COVID-19 deaths overall and by age in each period in 2022 in Hong Kong.**

| Age group | Observed deaths (Number) | Excess deaths (Number, 95% CI) | COVID-19 deaths (Number. 95% CI) | Differences between excess deaths and COVID-19 deaths |
|---|---|---|---|---|
| **All periods in 2022** | | | | |
| Overall | 63484 | 13518 (13402, 13633) | 12228 (12013, 12446) | 1290 |
| 0- | 86 | 19 (17, 22) | 9 (5, 17) | 10 |
| 5- | 47 | 0 (-2, 3) | 4 (2, 10) | -4 |
| 15- | 1328 | 293 (282, 305) | 100 (82, 122) | 193 |
| 45- | 8537 | 1082 (1048, 1115) | 846 (791, 905) | 236 |
| 65- | 17134 | 2959 (2903, 3015) | 2672 (2573, 2775) | 287 |
| 80- | 36352 | 9164 (9070, 9258) | 8597 (8419, 8779) | 567 |
| **Period 1 (weeks 1-26) in 2022** | | | | |
| Overall | 36848 | 10585 (10500, 10670) | 9738 (9547, 9933) | 847 |
| 0- | 41 | 6 (4, 8) | 5 (2, 12) | 1 |
| 5- | 25 | 2 (1, 4) | 2 (1, 7) | 0 |
| 15- | 666 | 116 (108, 125) | 77 (62, 96) | 39 |
| 45- | 4560 | 718 (694, 743) | 648 (600, 700) | 70 |
| 65- | 9502 | 2215 (2174, 2255) | 2103 (2015, 2195) | 112 |
| 80- | 22054 | 7527 (7457, 7597) | 6903 (6743, 7066) | 624 |
| **Period 2 (weeks 27-45) in 2022** | | | | |
| Overall | 18223 | 1341 (1278, 1404) | 1103 (1040, 1170) | 238 |
| 0- | 33 | 10 (8, 11) | 3 (1, 9) | 7 |
| 5- | 16 | 0 (-2, 1) | 1 (0, 6) | -1 |
| 15- | 488 | 128 (121, 135) | 12 (7, 21) | 116 |
| 45- | 2798 | 182 (163, 202) | 90 (73, 111) | 92 |

| | | | | |
|---|---|---|---|---|
| 65- | 5295 | 403 (372, 435) | 252 (223, 285) | 151 |
| 80- | 9593 | 617 (566, 668) | 745 (693, 800) | -128 |
| | | **Period 3 (weeks 46-52) in 2022** | | |
| Overall | 8413 | 1592 (1547, 1637) | 1387 (1316, 1462) | 205 |
| 0- | 12 | 3 (2, 4) | 1 (0, 6) | 2 |
| 5- | 6 | -2 (-3, -1) | 1 (0, 6) | -3 |
| 15- | 174 | 49 (45, 53) | 11 (6, 20) | 38 |
| 45- | 1179 | 181 (168, 194) | 108 (89, 130) | 73 |
| 65- | 2337 | 341 (319, 363) | 317 (284, 354) | 24 |
| 80- | 4705 | 1020 (983, 1056) | 949 (891, 1011) | 71 |

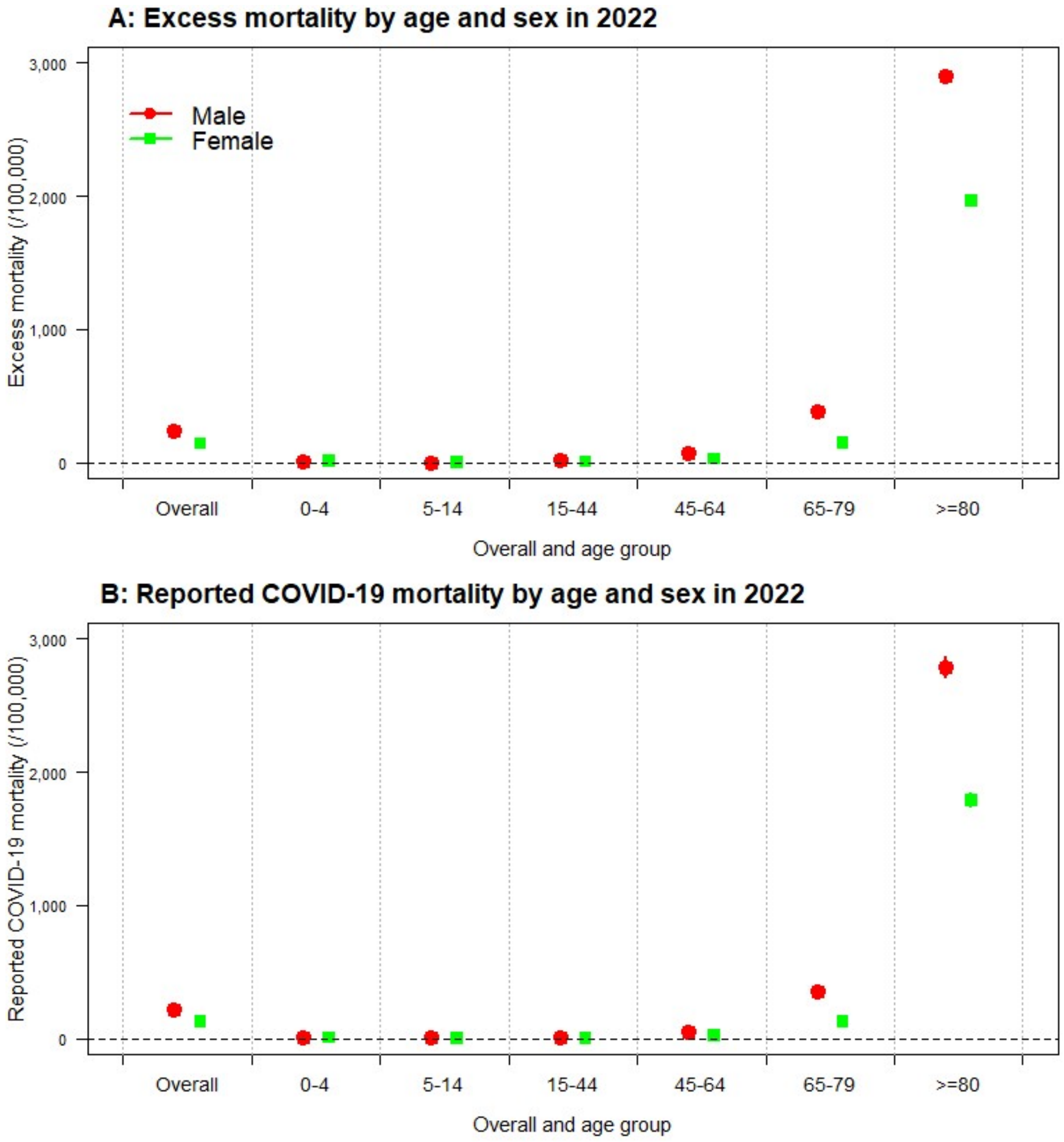


**Figure S1. Age- and sex-specific excess mortality (A) and reported COVID-19 mortality (B) in 2022.**

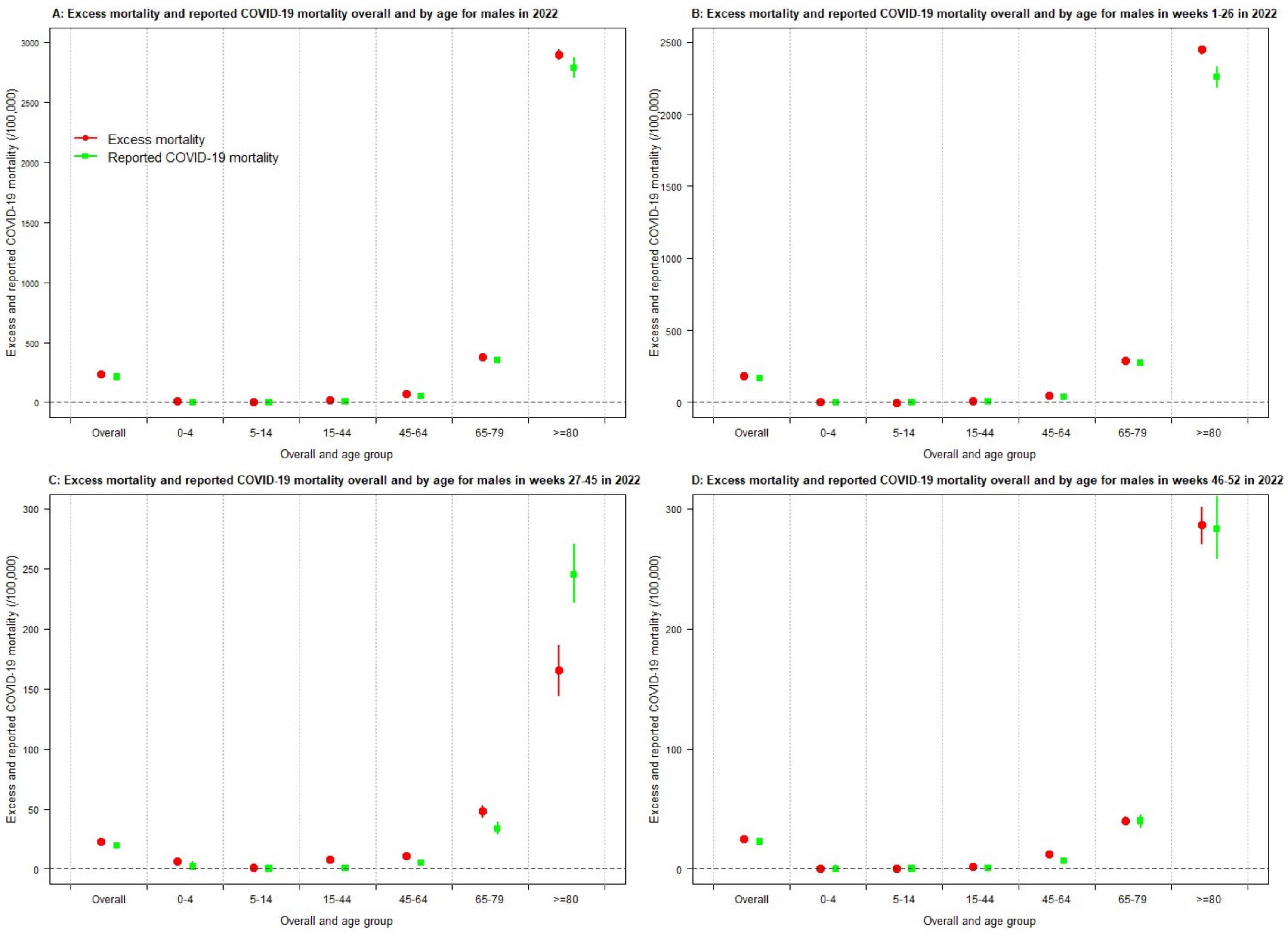


**Figure S2. Overall and age-specific excess mortality and reported COVID-19 mortality for males in 2022. A)** Comparison of excess mortality and reported COVID-19 mortality overall and by age for **males** in 2022. **B)** Comparison of excess mortality and reported COVID-19 mortality overall and by age for **males** in weeks 1-26 in 2022. **C)** Comparison of excess mortality and reported COVID-19 mortality overall and by age for **males** in weeks 27-45 in 2022. **D)** Comparison of excess mortality and reported COVID-19 mortality overall and by age for **males** in weeks 46-52 in 2022.

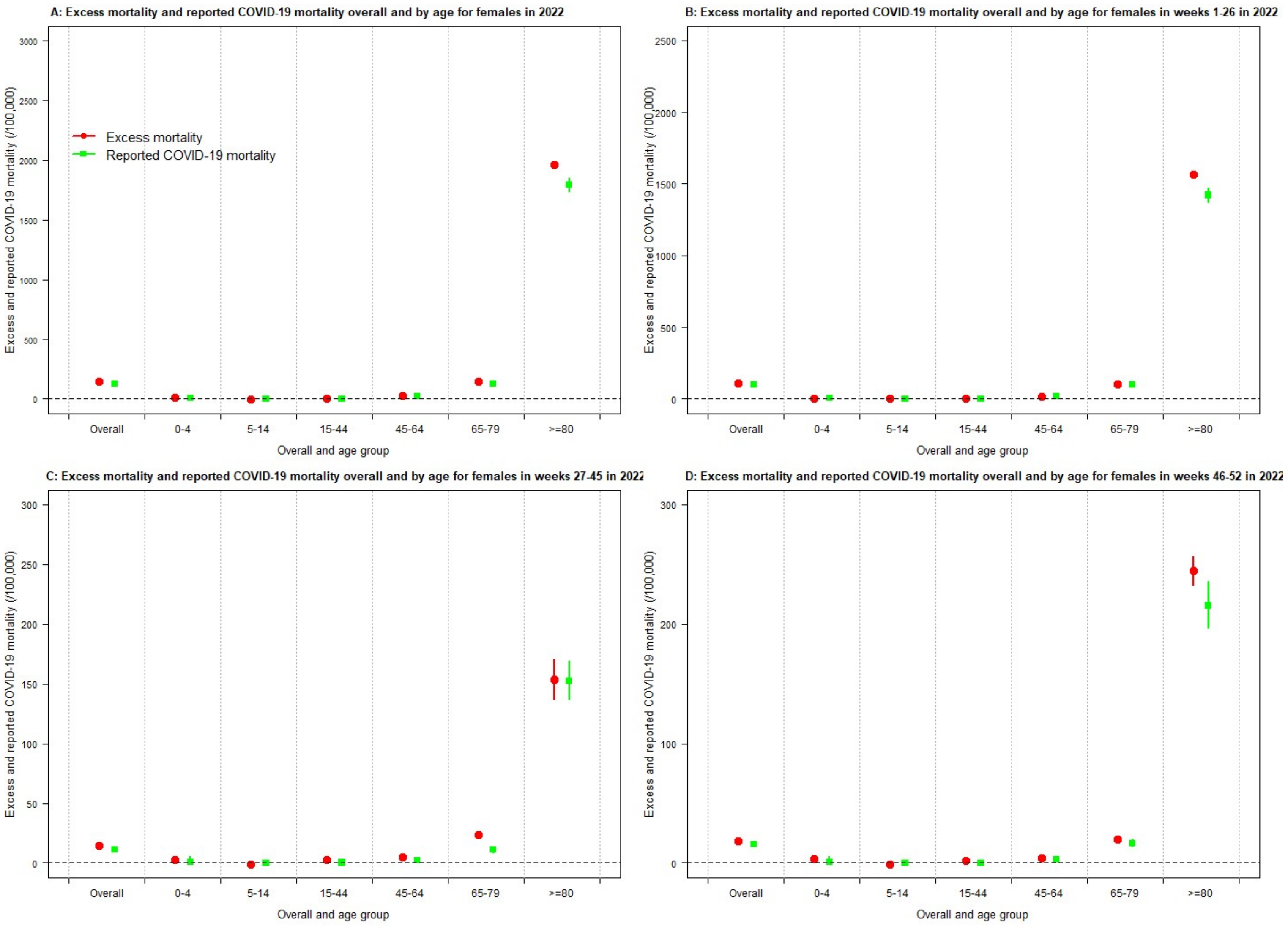


**Figure S3. Overall and age-specific excess mortality and reported COVID-19 mortality for females in 2022. A)** Comparison of excess mortality and reported COVID-19 mortality overall and by age for **females** in 2022. **B)** Comparison of excess mortality and reported COVID-19 mortality overall and by age for **females** in weeks 1-26 in 2022. **C)** Comparison of excess mortality and reported COVID-19 mortality overall and by age for **females** in weeks 27-45 in 2022. **D)** Comparison of excess mortality and reported COVID-19 mortality overall and by age for **females** in weeks 46-52 in 2022.

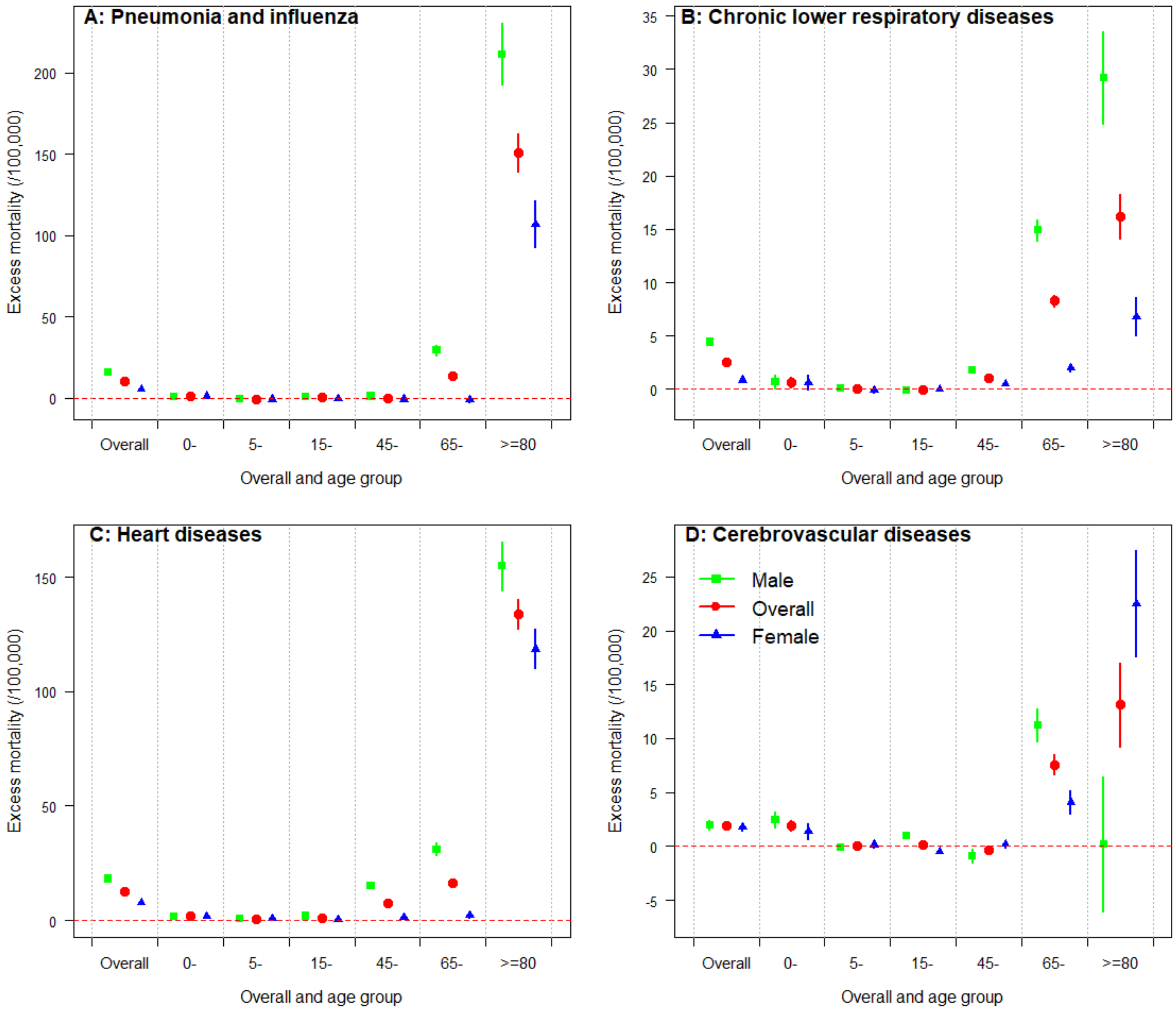


**Figure S4. Cause-specific excess mortality by age and sex in Hong Kong in 2022.**
A) Excess mortality by age and sex for Pneumonia and Influenza. B) Chronic lower respiratory diseases. C) Heart diseases. D) Cerebrovascular diseases.
